# Impact of Fluvoxamine on outpatient treatment of COVID-19 in Honduras

**DOI:** 10.1101/2022.09.27.22280428

**Authors:** Estela Pineda, Jarmanjeet Singh, Miguel Vargas Pineda, Jose Garay Umanzor, Fernando Baires, Luis G. Benitez, Cesar Burgos, Anupamjeet Kaur Sekhon, Nicole Crisp, Anita S. Lewis, Jana Radwanski, Marco Bermudez, Karen Sanchez Barjun, Oscar Diaz, Elsa Palou, Rossany E. Escalante, Carlos Isai Hernandez, Mark L. Stevens, Deke Eberhard, Manuel Sierra, Tito Alvarado, Omar Videa, Miguel Sierra-Hoffman, Fernando Valerio Pascua

**Affiliations:** Department of Internal Medicine Hospital CEMESA, San Pedro Sula, Cortes, Honduras; Department of Cardiovascular Medicine, University of California, Riverside, CA, United States of America; Department of Internal Medicine Hospital Mario Catarino Rivas, San Pedro Sula, Cortes, Honduras; Department of Obstetrics and Gynecology Hospital Mario Catarino Rivas, San Pedro Sula, Cortes, Honduras; Universidad Nacional Autónoma de Honduras, Tegucigalpa, Honduras; Sleep Medicine, Kaiser Permanente, Fontana, CA, United States of America; Wound Care Department El Campo Memorial Hospital, El Campo, TX, United States of America; Pharmacy Department El Campo Memorial Hospital, El Campo, TX, United States of America; Pharmacy Department Citizens Hospital, Victoria, TX, United States of America; Department of Medicine SBH Health System, Bronx, NY, United States of America; Department of Critical Care Hospital Regional del Norte Instituto Hondureño de Seguridad Social, San Pedro Sula, Cortés, Honduras; Internal Medicine Department, Facultad de Ciencias Médicas, Universidad Nacional Autónoma de Honduras, Tegucigalpa, Honduras; Department of Medicine, Facultad de Ciencas Médicas, Universidad Nacional Autónoma de Honduras, Tegucigalpa, Honduras; HEME Clinic, Choluteca, Choluteca, Honduras; Research Department, Texas A&M College of Medicine, Detar Family Medicine Residency Program, Victoria, TX, United States of America; Universidad Tecnológica Centroamericana, Tegucigalpa, Honduras; Infectiology Department, Facultad de Ciencias Médicas, Universidad Nacional Autónoma de Honduras, Tegucigalpa, Honduras; Clínica de Atención Medica Integral CAMI, Tegucigalpa, Honduras; Research and Infectious Disease Department, Texas A&M College of Medicine, Detar Family Medicine Residency Progra, Victoria, TX, United States of America; Department of Critical Care Hospital CEMESA, San Pedro Sula, Cortes, Honduras

**Keywords:** Fluvoxamine, COVID-19, Repurposed drugs, Early outpatient treatment, Honduras

## Abstract

**Background:** COVID-19 pandemic has impacted lives globally. While COVID-19 did not discriminate against developed or developing nations, it has been a significant challenge for third world countries like Honduras to have widespread availability of advanced therapies. The concept of early treatment was almost unheard-of when early outpatient treatment with repurposed drugs in Latin American countries showed promising results. One such drug is fluvoxamine, that has shown tremendous potential in two major studies, following which fluvoxamine was added to the standard of care in Honduras.

**Methods:** This is a prospective observational study performed at the Hospital Centro Médico Sanpedrano (CEMESA) in San Pedro Sula, Cortes, Honduras in the COVID-19 outpatient clinic. All patients fifteen years of age or older, with mild or moderate signs and symptoms of COVID-19, and a positive SARS-CoV-2 antigen or Reverse Transcription Polymerase Chain Reaction (RT-PCR) were included in the study and prescribed fluvoxamine. Cohort of patients who decided to take fluvoxamine were compared to the cohort who did not take fluvoxamine for mortality risk and risk of hospitalization as primary endpoints. Patient were monitored for 30 days with first follow up at 7 days and second follow up at 10-14 days of symptom onset. Categorical variables were compared by Pearson Chi-square test. The Odds ratio was calculated using univariate and multivariate logistic regression. Continuous variables were compared by t-test and Wilcoxon rank-sum tests.

**Results:** Of 657 total COVID-19 cases, 594 patients took fluvoxamine and 63 did not. A total of five patients (0.76 percent) died, of which only one death occurred in the fluvoxamine group. Patients who did not receive fluvoxamine had a significantly higher mortality (OR 24, p0.005, CI 2.6 to 233.5). Odds ratio of hospitalization in patients who did not take fluvoxamine was 2.38 (30 vs 10 hospitalizations, p 0.040, CI 1.04-5.47). The odds ratio of requiring oxygen in patients in the non-fluvoxamine group was 5.08 (p<0.001, CI 2.18-11.81). Mean lymphocytes count on the first follow-up visit was significantly higher in the fluvoxamine group (1.72 vs. 1.38, Δ 0.33, p 0.007, CI 0.09 to 0.58).

**Conclusion:** The results of our study suggest lowers odds of mortality and hospitalization in patients who took fluvoxamine vs fluvoxamine non-takers. Non-fluvoxamine group had higher odds of oxygen requirement than fluvoxamine group as well.

## Introduction

The coronavirus disease 2019 (COVID-19) pandemic that originated in Wuhan, China remains not only a topic of debate but also poses a threat to our healthcare systems, society, and overall economy. For most of 2020 and part of 2021, skepticism, doubt, and fear of the unknown could be globally palpated. The concept of early treatment was almost unheard-of; instead, most international scientific bodies only recommended isolating at home and symptomatic treatment. As a result, due to lack of consistent therapeutic guidelines many patients were left to anxiously wait, developing severe disease before a healthcare provider would assess their condition. At this point, it was often too late to prevent them from succumbing to the virus.

As early as January 2020, the clinical evidence became clear and patients with Severe Acute Respiratory Syndrome Coronavirus 2 (SARS-CoV-2) demonstrated an identifiable pattern. Survivors were young and typically showed very low or minimal activated inflammatory markers and thrombotic state. Non-survivors, on the other hand had quite the opposite profile with hyperinflammatory and pro-coagulant immunological profiles (JAMA 2020).

Even with this evidence, early outpatient treatment with repurposed drugs was not embraced by most developed nations (Valerio et al., 2022). However, in Latin American countries, early treatment was adopted soon after early observational studies in these patients showed promising results (Ontai et al, 2022).

One treatment in particular, fluvoxamine, a first-generation selective serotonin reuptake inhibitor (SSRI), showed tremendous potential in not only one but two studies. (Reis et al., 2022; Seftel and Boulware, 2021). Interestingly, beside the more traditional antidepressant and anxiolytic properties of SSRIs these family of drugs display anti-inflammatory properties that could be of potential use in ameliorating the hyperinflammatory phases of the disease (Meikle et al., 2021).Even though Honduras is a low to middle income Central American country with an extremely underserved and understaffed medical system, the government proactively recommended early outpatient treatment of COVID-19 to avoid a collapse of their healthcare system. By the spring of 2020, based on the available evidence, safe repurposed drugs were already being used in an established protocol resulting in a significant reduction of the case fatality rate, only one week after initiation of this national protocol (Ontai et al., 2022; Secretaría de Salud De Honduras 2020; Valerio et al., 2021). After the publication of a randomized control study by Lenze et al. that showed prevention of clinical deterioration in the early treatment for COVID-19 with fluvoxamine use (Lenze et al., 2020); fluvoxamine was offered as a treatment option in addition to the standard of care protocol in Honduras (Valerio et al., 2022). Here we present a real-world observational study done in Honduras, a nation in development to evaluate the effectiveness of fluvoxamine in preventing clinical deterioration in terms of mortality and hospitalization.

## Methods

We conducted this prospective observational study with data collected from medical records from November 2020 until January 2022. The study was approved by Institutional Review Board (IRB) of the Hospital Centro Médico Sanpedrano and the Ethics committee of investigation of infectious and zoonotic disease masters of the Universidad Nacional Autónoma de Honduras. This study was performed in the COVID-19 outpatient clinic at the Hospital Centro Médico Sanpedrano (CEMESA) in San Pedro Sula, Cortes, Honduras. Patients fifteen years of age or older, with mild to moderate COVID-19 with a positive SARS-CoV-2 antigen or Reverse Transcription Polymerase Chain Reaction (RT-PCR) were included in the study. Patients who developed signs and symptoms of the disease and were in close contact with one of the patients in the cohort were also treated and included in the study. Pregnant women, patients younger than fifteen years of age, and patients with severe and critical COVID-19 presentation were excluded.

Fluvoxamine was prescribed as part of the early treatment to all patients presenting with mild to moderate COVID-19 disease. While fluvoxamine was recommended to everyone, a cohort of patients chose not to take fluvoxamine and became our control group. All patients were monitored for 30 days and had a follow up evaluation at 7 and 10-14 days after symptom onset. Patients were followed using telemedicine if they had new concerning symptoms such shortness of breath, chest pain, or oxygen desaturation. Patients were started on fluvoxamine 50 mg orally twice daily for three days and titrated up to 100 mg two or three times a day depending on patient tolerance and disease severity to complete a fourteen-day course (Valerio et al., 2022).

Mild Disease: Patients presenting with any of the common signs or symptoms of SARS-CoV-2 infection, including fever, cough, sore throat, malaise, headache, myalgias, nausea, vomiting, diarrhea or loss of taste and smell, they do not have dyspnea, hypoxia, or abnormal chest radiography.

Moderate Disease: Patients who show evidence of lower respiratory disease (e.g., dyspnea) at clinical presentation, and/or abnormal chest radiography, and who have a pulse oximetry of ≥ 94% on room air.

Severe Disease: Patients who show evidence of lower respiratory disease with a pulse oximetry of < 94% on room air, a ratio of arterial partial pressure of oxygen to fraction of inspired oxygen (PaO_2_/FiO_2_) < 300 mmHg, a respiratory rate of > 30 breaths per min, and/or abnormal chest radiography demonstrating > 50% pulmonary infiltrates.

Critical Illness: Individuals presenting with respiratory failure, septic shock, and/or multiple organ dysfunction.

### Statistical analysis

Statistical analysis was performed using Stata 17.0 Basic edition software. Baseline descriptive statistics were performed on all variables. Inferential statistics were conducted via Pearson Chi-square testing for categorical data outcomes. The Odds ratio was calculated using univariate logistic regression. We then performed multivariate logistic regression by modelling only significant variables from the univariate logistic regression model. Continuous variables were compared by t-test and Wilcoxon rank-sum tests. Wilcoxon rank-sum test was used primarily for non-parsimonious continuous data variables.

## Results

Of 657 total patients in the study, 330 were males (50.2%). 595 patients took fluvoxamine, and 62 patients did not take fluvoxamine. Mean age of fluvoxamine group was 47.9 years and of non-fluvoxamine group was 49.7 years (p value 0.409). A total of 170 patients had hypertension (158 in fluvoxamine group vs 12 in non-fluvoxamine group, p 0.218), 88 had Diabetes (79 in fluvoxamine group vs 9 in non-fluvoxamine group, p 0.785), and 202 were obese (185 in fluvoxamine group vs 17 in non-fluvoxamine group, p 0.697). 243 patients had complete vaccination whereas 115 had incomplete vaccination and 299 were unvaccinated. (Figure 1) (Table 1)

**Table 1.** Baseline characteristics of the study cohort according to fluvoxamine use.

| *Characteristic* | **Cohort**<br>**Participants** | **Received Fluvoxamine** | **Not- received fluvoxamine** | **P Value** |
| --- | --- | --- | --- | --- |
|  | No (%) | no. | no. |  |
| **Total** | 657 | 594 (90.6 %) | 63 (9.4 %) |  |
| **Sex** |  |  |  | 0.761 |
| *Female* | 327 (49.8) | 295 (44.9 %) | 32 (4.9 %) |  |
| *Male* | 330 (50.2) | 300 (45.7 %) | 30 (4.6 %) |  |
| **Mean Age** | 48.1 years | 47.9 years | 49.7 years | 0.409 |
| **Comorbidities** |  |  |  |  |
| *Hypertension* | 170 (25.8) | 158 (24.1 %) | 12 (1.9 %) | 0.218 |
| *Diabetes* | 88 (13.4) | 79 (12.0 %) | 9 (1.4 %) | 0.785 |
| *Obesity* | 202 (32.7) | 185 (28.2 %) | 17 (2.6 %) | 0.697 |
| *Dyslipidemia* | 29 (4.4) | 29 (4.4 %) | 0 (0.0 %) | 0.075 |
| *Autoimmune* | 36 (5.5) | 34 (5.2 %) | 2 (0.3 %) | 0.413 |
| **Vaccination Status** |  |  |  |  |
| *Complete* | 245 (37.3) | 240 (36.5 %) | 3 (0.5 %) | <0.001 |
| *Incomplete* | 112 (17.0) | 112 (17.1 %) | 3 (0.5 %) | 0.006 |
| *Unvaccinated* | 300 (45.8) | 243 (40.1 %) | 56 (8.5 %) | <0.001 |
| **Symptoms** |  |  |  |  |
| *Fever* | 195 (29.6) | 173 (26.3 %) | 22 (3.3 %) | 0.425 |
| *Arthralgia* | 111 (16.9) | 104 (15.9 %) | 7 (1.1 %) | 0.150 |
| *Myalgia* | 139 (21.1) | 126 (19.2 %) | 13 (19.8 %) | 0.869 |
| *Chest Tightness* | 71 (10.8) | 65 (9.9 %) | 6 (0.9 %) | 0.755 |
| *Headache* | 191 (29.1) | 173 (26.3%) | 18 (2.7 %) | 0.927 |
| *Anosmia* | 157 (23.9) | 144 (22.1 %) | 13 (1.9 %) | 0.455 |
| *Ageusia* | 131 (19.9) | 121 (18.4 %) | 10 (1.5 %) | 0.329 |
| *Odynophagia* | 122 (18.5) | 107 (16.3 %) | 15 (2.9 %) | 0.301 |
| *Dysgeusia* | 34 (5.2) | 31 (4.7 %) | 3 (0.5 %) | 0.997 |
| *Rhinorrhea/cong* | 151 (22.9) | 141 (21.5 %) | 10 (1.5 %) | 0.207 |
| *Hyporexia* | 122 (18.5) | 116 (17.6 %) | 6 (0.9 %) | 0.055 |
| *Anorexia* | 36 (5.5) | 31 (4.7 %) | 5 (0.7 %) | 0.291 |
| *Insomnia* | 77 (11.7) | 72 (10.9 %) | 5 (0.7 %) | 0.358 |
| *Cough* | 187 (28.5) | 174 (26.5 %) | 13 (19.8 %) | 0.147 |
| *Shortness of Breath* | 70 (10.6) | 57 (8.7 %) | 13 (19.8 %) | 0.003 |
| *Expectoration* | 49 (7.5) | 43 (6.5 %) | 6 (0.9 %) | 0.614 |
| *Diarrhea* | 102 (15.5) | 94 (14.3 %) | 8 (1.2 %) | 0.972 |
| *Malaise* | 213 (32.4) | 189 (28.8 %) | 24 (36.5 %) | 0.297 |
| *Nausea* | 6 (0.9) | 6 (0.9 %) | 0 (0.0 %) | -- |
| *Vomiting* | 2 (0.3) | 2 (0.3 %) | 0 (0.0 %) | -- |
| **Initial Covid Stage** |  |  |  | 0.001 |
| *Mild* | 554 (84.3) | 497 (75.6 %) | 57 (8.7 %) |  |
| *Moderate* | 102 (15.5) | 98 (15.0 %) | 4 (0.6 %) |  |
| *Severe* | 1 (0.15) | 0 (0.0 %) | 1 (0.2 %) |  |

**Figure 1.**
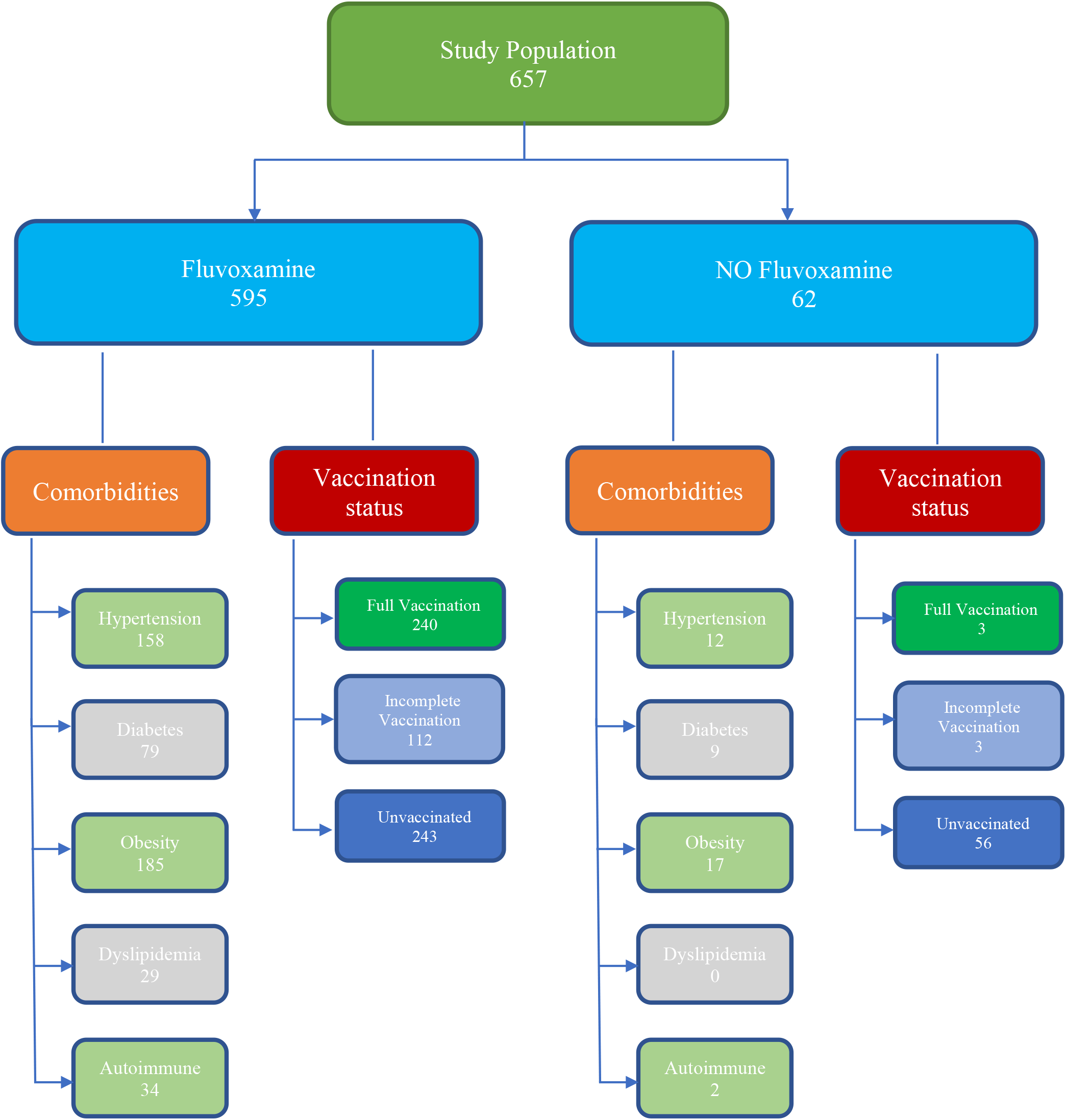

### 1. Primary Endpoints

#### 1.1. 30-day-Mortality

Of 657 total COVID-19 cases, 5 patients (0.76%) died. Only one death occurred in the fluvoxamine group. Pearson chi-square test showed a significant association between lack of fluvoxamine use and mortality (χ^2^ = 28.8, p <0.001, Cramer’s V 0.21). Patients who did not receive fluvoxamine had a significantly higher 30-day mortality risk (OR 40, p0.001, CI 4.4 to 365). The odds ratio for mortality in the non-fluvoxamine group remained significant despite controlling for hypertension, diabetes, age, vaccination status, or premedication with steroids (OR 24, p 0.005, 95%CI 2.6 to 233.5). (Table 2)

**Table 2.** Results: Primary and Secondary Outcomes

Results: Primary and Secondary Outcomes
| *Outcomes* | **Total no.** | **Received**<br>**Fluvoxamine** | **Not- received**<br>**fluvoxamine** | **Odds Ratio, P Value, 95% CI** |
| --- | --- | --- | --- | --- |
|  | Total | No (%) | No (%) |  |
| *Study population* | 657 | 595 (90.6) | 62 (9.4) |  |
| *Primary outcomes* |  |  |  |  |
| *Death* | 5 | 1 (20.0) | 4 (80.0) | OR 24 , p 0.005, CI 2.6 to 233.5 |
| *Hospitalization* | 40 | 30 (5% of 595) | 10 (15% of 63) | OR 2.38, p 0.040, CI 1.04-5.47 |
| *Secondary outcomes* |  |  |  |  |
| *Oxygen requirement* | 33 | 21/595 (3.5) | 12/62 (19.4) | OR 5.08, p<0.001, CI 2.18-11.81 |
| *Tocilizumab* | 10 | 6/595 (1) | 4/62 (6.5) | OR 2.67, p 0.215, CI 0.56 -12.55 |
| *Days in hospital* | | 10.3 days | 7.6 days | $\Delta$ 2.7, p 0.328, CI -8.1 to 2.8 |
| *Mean Lymphocyte count* | | 1.72 (10<sup>3</sup>/ul) | 1.38 (10<sup>3</sup>/ul) | $\Delta$ 0.33, p 0.007, CI 0.09 to 0.58 |

#### 1.2. Hospitalization

Out of 657 total patients in the study, 40 (6%) were hospitalized with COVID-19. 5% (30 out of 594) of patients who received fluvoxamine and 15% (10 out of 63) of patients who did not take fluvoxamine were hospitalized. There was a significant difference in hospitalization between two groups (Pearson χ^2^ 12.07, p 0.001). Odds ratio of hospitalization in patients who did not take fluvoxamine was 3.62 (p 0.001, 95%CI 1.67-7.82) versus who took fluvoxamine. Odds of being hospitalized in non-fluvoxamine group remained statistically significant despite adjusting for age, hypertension, diabetes, and vaccination status (OR 2.38, p 0.040, 95%CI 1.04-5.47) (Table 2)

### 2. Secondary Endpoints

#### 2.1 Oxygen requirement

A total of 30 patients required oxygen. Of those, 24 were hospitalized and 9 were in non-hospitalized group. Mean age of patients that required oxygen was significantly higher (55.5 vs 47.7, p=0.007). The Odds ratio of requiring oxygen in patients in non-fluvoxamine group after controlling for hypertension, diabetes, age, and vaccination status was 5.08 (p<0.001, CI 2.18-11.81) (Table 2)

#### 2.2 Tocilizumab requirement

Of 657 total patients, a total of 10 patients required tocilizumab. Odds ratio of requiring tocilizumab in patients who did not take fluvoxamine was 6.77 (p 0.004, CI 1.85-24.68) versus who took fluvoxamine. However, when stratified by hospitalization, the odds ratio of requiring tocilizumab failed to reach significance between two groups (OR 2.67, p 0.215, CI 0.56 -12.55). (Table 2)

#### 2.3 Days in hospital

Mean hospital stay for hospitalized patients was 8.3 days (median 6 days). There was no significant difference in the mean hospital stay between hospitalized patients from fluvoxamine and non-fluvoxamine groups (10.3 vs 7.6 days, p0.328). (Table 2)

#### 2.4 Critical Illness

Out of total 657 cases, 624 (95.0 percent) stayed on mild to moderate COVID 19 disease, and 33 (5.0 percent) advanced to severe to critical disease. Fluvoxamine group had significantly lower number of cases advancing to a severe-to-critical COVID-19 stage than non-fluvoxamine group (Pearson χ^2^ 43.2, p <0.001, Cramer’s V 0.26)

#### 2.5 Laboratory Markers

Mean lymphocytes count on the first follow up visit was significantly higher in fluvoxamine group (1.72 vs 1.38, Δ 0.33, p <0.006, 95%CI 0.09 to 0.58). Mean WBC count on the first follow up visit was significantly different between non-fluvoxamine and fluvoxamine group (7.8 vs 5.9, Δ 1.9, p <0.001, CI 0.9 to 2.9). Mean CRP levels were not statistically different between non-fluvoxamine and fluvoxamine group (30.3 vs 19.4, Δ 10.9, p 0.216, CI 6.4 to 28.1). Similarly, mean procalcitonin, d-dimer, and serum ferritin levels were not statistically different between non-fluvoxamine and fluvoxamine group (p >0.05). (Table 2)

## Discussion

Multiple mechanisms have been described in which fluvoxamine can prevent deterioration and hospitalization in patients with multiple risk factors for COVID-19 disease progression. SSRIs have been shown to reduce serum serotonin levels by >80% (Duerschmied et al., 2013). Declining levels of serum serotonin reduce platelet aggregation and increase bleeding time (Halperin et al., 2007). In hospitalized COVID-19 patients with acute respiratory distress syndrome (ARDS), there has been described platelet activation hypersensitivity compared to non-COVID-19 ARDS patients (Zaid et al., 2021). This most likely is derived from excessive free plasma serotonin levels from virus induced excessive platelet activation, that leads to excessive platelet-fibrin aggregation and formation of microthrombi. This contributes to clinical deterioration, a mechanism which fluvoxamine could block through its serotonin reducing effects on plasma and anti-aggregation effects on platelets. Fluvoxamine has also been shown to play a significant role in modulating inflammation through its agonistic activity on the sigma-1 receptor. Through its chaperone activity, it may protect against mitochondrial damage and endoplasmic reticulum (ER) stress in response to SARS-COV2 infection, as it prevents protein misfolding in the ER because of the ER overloading with virus-encoded proteins (Hayashi et al., 2007; Brimson et al., 2021). The sigma-1 receptor chaperone activity is stimulated through the unfolded protein response (UPR), triggered by ER stress, in which the UPR facilitates proper protein folding within the endoplasmic reticulum, preventing cellular stress on the endoplasmic reticulum. If cellular stress on ER is unbearable, it can lead to autophagy and inflammation that can trigger a cytokine storm (Fung et al., 2014). The sigma-1 receptor has also been shown to be essential in modulating inflammatory reactions in pre-clinical studies, in which mice lacking the sigma-1 receptor (through knock out gene modification) that were injected with lipopolysaccharides (LPS), had higher levels of Tumor necrosis factor (TNF) and Interleukin-6 (IL-6). This contributed to more severe sepsis compared to the control mice group injected with LPS with intact sigma-1 receptors (Rosen et al., 2019). Fluvoxamine has been shown to attenuate inflammation in rat models with experimental autoimmune encephalomyelitis (Ghareghani et al., 2017). This could be explained by the fact that fluvoxamine is a potent agonist of the sigma-1 receptor, which could give a mechanism for modulation of inflammation through its sigma-1 receptor chaperone activity. Such agonist action could prevent the robust cytokine release phenomena with subsequent exuberant inflammatory response, thus preventing clinical deterioration of COVID-19 ambulatory patients with mild symptoms at risk of progression to severe disease and hospitalization.

Other mechanisms of fluvoxamine that could account for its efficacy on prevention of hospitalization of mild COVID-19 ambulatory cases could be its inhibitory effect on acid sphingomyelinase (ASM). This results in reduced ceramide-enriched membrane domains on the membrane of epithelial cells. These are the domains SARS-COV-2 uses for facilitation of entry through clustering of acetylcholine esterase 2 (ACE2) receptors, the main cellular receptor of SARS-COV-2. The SARS-CoV-2 virus stimulates the ASM/ceramide system, producing ceramide rich domains that cluster the ACE2 receptors, then facilitates the entry of the SARS-CoV-2 virus into the epithelial cells. Fluvoxamine can block this mechanism of infection through inhibition of the ASM, preventing entry of the virus into cells, and thus diminishing viral load in patients therefore preventing infection, reducing virulence, and reducing risk of severe disease (Kornhuber et al., 2021). Fluvoxamine also increases melatonin levels in patients through inhibition of cytochrome P450 enzyme CYP1A2. This can potentiate the anti-inflammatory, antioxidant, and immunomodulatory mechanisms melatonin has on various viral infections, including COVID-19 (Anderson et al., 2020). SSRIs like fluvoxamine also decrease histamine release from mast cells (Ferjan and Erjavec, 1996). Histamine plays a vital role in inflammation, edema, and thrombosis in patients hospitalized by COVID-19. Plasma levels of chymase, a serum marker of mast cell degranulation, were significantly more elevated in hospitalized COVID-19 patients compared to community ambulatory cases (Tan et al., 2021). This elevation signifies the important role mast cells have in the cytokine release phenomena with subsequent exuberant inflammatory response in hospitalized COVID-19 patients. Their role is demonstrated in postmortem COVID-19 patients’ lung biopsies in which activated macrophages were linked to pulmonary edema and thrombosis (Motta Junior et al., 2020). All these mechanisms suggest fluvoxamine has significantly more benefits than its psychotropic effects and can have a critical role in treatment of COVID-19 patients.

The TOGETHER trial showed an absolute risk reduction of 5·0%, and 32% relative risk reduction, on the primary outcome of hospitalization, with fluvoxamine administration for 10 days (Reis et al., 2022) Additionally, these results support those of Lenze et al that were replicated with another real-world publication by Seftel et al. (Lenze et al., 2020; Seftel and Boulware, 2021) In addition, the findings of this study include the benefits of this drug among vaccinated people with high risk for clinical deterioration, who despite the protection of the vaccines, often are reported with severe disease.

The discussed anti-inflammatory, hemostatic, anti-viral, and antihistamine properties of fluvoxamine can explain the significant difference of lower hospitalizations and mortality in the treatment group compared to the placebo group as shown in our study.

### Limitations

Prospective cohort studies, being non-randomized, carry inherent biases related to lack of randomization and confounding. However, our study is real world, involving a large cohort of patients, and has statistical adjustment for various factors with multivariate analysis. Secondly, this study was performed in the outpatient setting. As a result, close hemodynamic monitoring and care of patients was not performed in the hospital. The difference in outcomes with fluvoxamine could be affected by social support and medication compliance. However, our patients received the standard COVID -19 protocol in both arms and followed up with us as per standard study protocol. The lack of ethnic diversity, and the fact that it was conducted in a single hospital during the COVID-19 pandemic are all limiting factors with respect to the study outcomes. The study was conducted at a private hospital, some patients chose to be hospitalized even if they did not meet hospitalization criteria and increasing the hospitalization rate. Despite all these limitations, there’s a counterbalance when using fluvoxamine in an underdeveloped nation like the one doing the study. The pandemic has proven to the world that there have been disparities in the distribution of novel therapies directed to SarsCoV2, in the vaccines and monoclonal antibodies. Given our prior experiences in the distribution of new therapies, it would be naïve to believe that Honduras will be a priority to these big pharmaceutical companies producing these products.

### Social and Economic Impact

Fluvoxamine represents a relatively inexpensive option that is affordable for people who live in extreme poverty in developing countries such as Honduras. Fluvoxamine has already been approved and discussed in most pharmacies around the world, therefore, is more accessible including rural areas which will have a bigger impact in the developing nations. The evidence that we presented in this study will have both social and economic impact on the ambulatory management of COVID-19 patients.

## Conclusions

Despite new antiviral medications, such as Molnupiravir and Paxlovid, showing promise in the prevention of disease severity, the production and distribution of these drugs to underserved people in low to middle income countries such Honduras will be limited due to the high demand of these drugs in developed countries. Additionally, the high selling prices of these drugs will limit government investment in these countries. Furthermore, the benefit of fluvoxamine goes beyond its antiviral potential and delivers fundamental benefits by reducing the impact on the mechanisms of disease severity that are associated with death. The study showed a significant mortality and hospitalization benefit which was similar to the results of a recent publication, the TOGETHER trial, among patients who completed treatment with fluvoxamine.

## Data Availability

All data produced in the present work are contained in the manuscript

## Acknowledgments

We wish to thank pharmacists Mr. Antonio Handal Mourra, and Mr. Jose Francisco Simán Benedetto for their altruism and constant support and dedication with this study and during the pandemic in Honduras.

## Notes

### Competing Interest Statement

The authors have declared no competing interest.

### Funding Statement

This study did not receive any funding

### Author Declarations

Institutional Review Board (IRB) of the Hospital Centro Medico Sanpedrano and the Ethics committee of investigation of infectious and zoonotic disease masters of the Universidad Nacional Autonoma de Honduras gave ethical approval for this work.

